# Consensus gene modules strategy identifies candidate blood-based biomarkers for primary Sjögren’s disease

**DOI:** 10.1101/2023.07.05.23292036

**Authors:** Cheïma Boudjeniba, Perrine Soret, Diana Trutschel, Antoine Hamon, Valentin Baloche, Bastien Chassagnol, Emiko Desvaux, Antoine Bichat, Audrey Aussy, Philippe Moingeon, Céline Lefebvre, Sandra Hubert, Marta Alarcón-Riquelme, Wan-Fai Ng, Jacques-Eric Gottenberg, Benno Schwikowski, Michele Bombardieri, Joel A.G. van Roon, Xavier Mariette, Mickaël Guedj, Etienne Birmele, Laurence Laigle, Etienne Becht

## Abstract

Primary Sjögren disease (pSD) is an autoimmune disease characterized by lymphoid infiltration of exocrine glands leading to dryness of the mucosal surfaces and by the production of autoantibodies. The pathophysiology of pSD remains elusive and no treatment with demonstrated efficacy is available yet. To better understand the biology underlying pSD heterogeneity, we aimed at identifying Consensus gene Modules (CMs) that summarize the high-dimensional transcriptomic data of whole blood samples in pSD patients. We performed unsupervised gene classification on four data sets and identified thirteen CMs. We annotated and interpreted each of these CMs as corresponding to cell type abundances or biological functions by using gene set enrichment analyses and transcriptomic profiles of sorted blood cell subsets. Correlation with independently measured cell type abundances by flow cytometry confirmed these annotations. We used these CMs to reconcile previously proposed patient stratifications of pSD. Importantly, we showed that the expression of modules representing lymphocytes and erythrocytes before treatment initiation is associated with response to hydroxychloroquine and leflunomide combination therapy in a clinical trial. These consensus modules will help the identification and translation of blood-based predictive biomarkers for the treatment of pSD.

## Introduction

Primary Sjögren Disease (pSD) is a chronic, disabling inflammatory autoimmune disease characterized by lymphoid infiltration of exocrine glands leading to dryness of the mucosal surfaces, such as the mouth and eyes and by the production of specific auto-antibodies[1–3]. Long-term complications include ocular and dental diseases, systemic involvement, organ damages and increased risk of lymphoma with excess mortality[4, 5]. This pathology is affecting between 0.05% and 0.4% of the adult population[6–9] and is the second most common systemic autoimmune disease[10]. It affects women more often than men (9:1) and the peak frequency of the disease is around fifty years of age[11].

The advent of new technologies has provided a path towards the development of classification criteria for autoimmune diseases that are based on molecular patterns representing disease mechanisms and molecular pathways[12, 13]. By applying computational methodologies to clinical and multiomic datasets, several pSD disease taxonomies have recently been proposed. Indeed, Tarn et al. proposed a symptom-based stratification of patients with pSD[14], while Soret et al.[15] and Trutschel et al.[16] proposed a molecular classification of pSD based on whole blood transcriptomic profiles of pSD patients. These classifications may provide useful clinical insights on disease subtypes of pSD patients but remain limited in the characterization of the biology underlying the disease in each patient subgroup. Indeed, pathogenesis of autoimmunity involves dysfunction of the entire immune system, and many cellular or functional components, including neutrophils, dendritic cells (DCs), macrophages, T and B cells, cytokine signaling pathways or autoantibodies[17, 18].

The clinical manifestations and biological disturbances associated with pSD are indeed highly heterogeneous among individuals which complicates its diagnosis. Mechanistically, the pathophysiology of pSD remains elusive[19]. No targeted therapy is therefore currently approved and only symptomatic treatments are offered[20, 21]. Precision Medicine approaches designed to better address the needs of patients based on the specific biological mechanisms underlying their symptoms would greatly improve the management of patients suffering from pSD.

The IMI 2 NECESSITY European consortium was launched in 2019 to identify a new composite clinical endpoint, biomarkers for stratifying patients and predictive biomarkers of treatment response for pSD, and test them in a prospective clinical trial. To achieve these goals, members of the NECESSITY consortium share clinically-annotated datasets, including whole blood transcriptomic datasets of pSD patients. These transcriptomes allow the identification of biological heterogeneity across pSD patients and its potential link with response to treatments, but were produced using diverse transcriptomic technologies, making their combined analysis challenging.

In order to jointly analyze independent whole blood transcriptomic datasets of pSD patients, we used a graph theoretical approach to unify four correlation networks into a consensus graph linking positively correlated genes. By clustering this unified representation of multiple cohorts, we identified 13 consensus transcriptomic gene modules that summarize the pathophysiology of pSD at the blood level. We annotated each of these modules for correspondence with cell types or molecular pathways, and validated these biological interpretation with matching flow cytometry data or cytokine measurements whenever available. We used these modules to better characterize and reconcile previously-published pSD patient stratifications[15, 16]. Importantly, we investigated clinical trial data to decipher the impacts of treatments on the peripheral blood of patients and propose a model predictive of the response to leflunomide-hydroxychloroquine combination therapy.

## Results

### Identification of thirteen consensus gene modules (CMs) from whole blood transcriptomes of pSD patients

We analyzed four whole blood transcriptomic datasets from pSD patients. Three were provided by the NECESSITY consortium: ASSESS[22] (*n* = 371), PreciseSADS[12] (*n* = 341) and UKPSSR[23] (*n* = 144). We also included the publicly-available GSE84844[24] dataset (*n* = 30). Our goal was to identify consistent signals across these four sources, and in particular consensus gene modules (CMs) of coexpressed genes. Transcriptomic data sets are however high dimensional which can hamper the correct identification of gene modules. Indeed, spurious correlations may appear due to the size and noisiness of the data: 20,000 protein coding genes indeed correspond to 400 *×* 10^6^ correlation coefficients. To ensure that the CMs we identify were reproducible across a large range of blood transcriptomic data sets (from distinct pSD cohorts), we used a dedicated analysis workflow summarized in **Figure 1A**. We first converted each cohort’s gene expression matrix to an affinity matrix (gene co-expression network). This affinity is non-linearly and monotonically linked to the observed correlation between two genes and shrinks low correlation coefficients towards 0 (See **Methods** and Wang *et al*.[25]). We applied Similarity Network Fusion (SNF)[25], a computational method designed for the merging of multiple affinity matrices, generating a consensual representation of genes’ pairwise similarities in the blood of pSD patients across these four independent cohorts (**Figure 1B**). We pruned the consensual affinity matrix to obtain a sparse weighted graph with edges corresponding to highly co-expressed genes (**Supplementary Figure 1**). Finally, Louvain clustering[26] of the sparse graph (see **Methods**) identified 13 CMs (**Supplementary Table 1**). We confirmed a posteriori that these CMs are reproducible groups of highly co-expressed genes that are reproducible across the four datasets (**Figure 1C**).

**Fig. 1.**
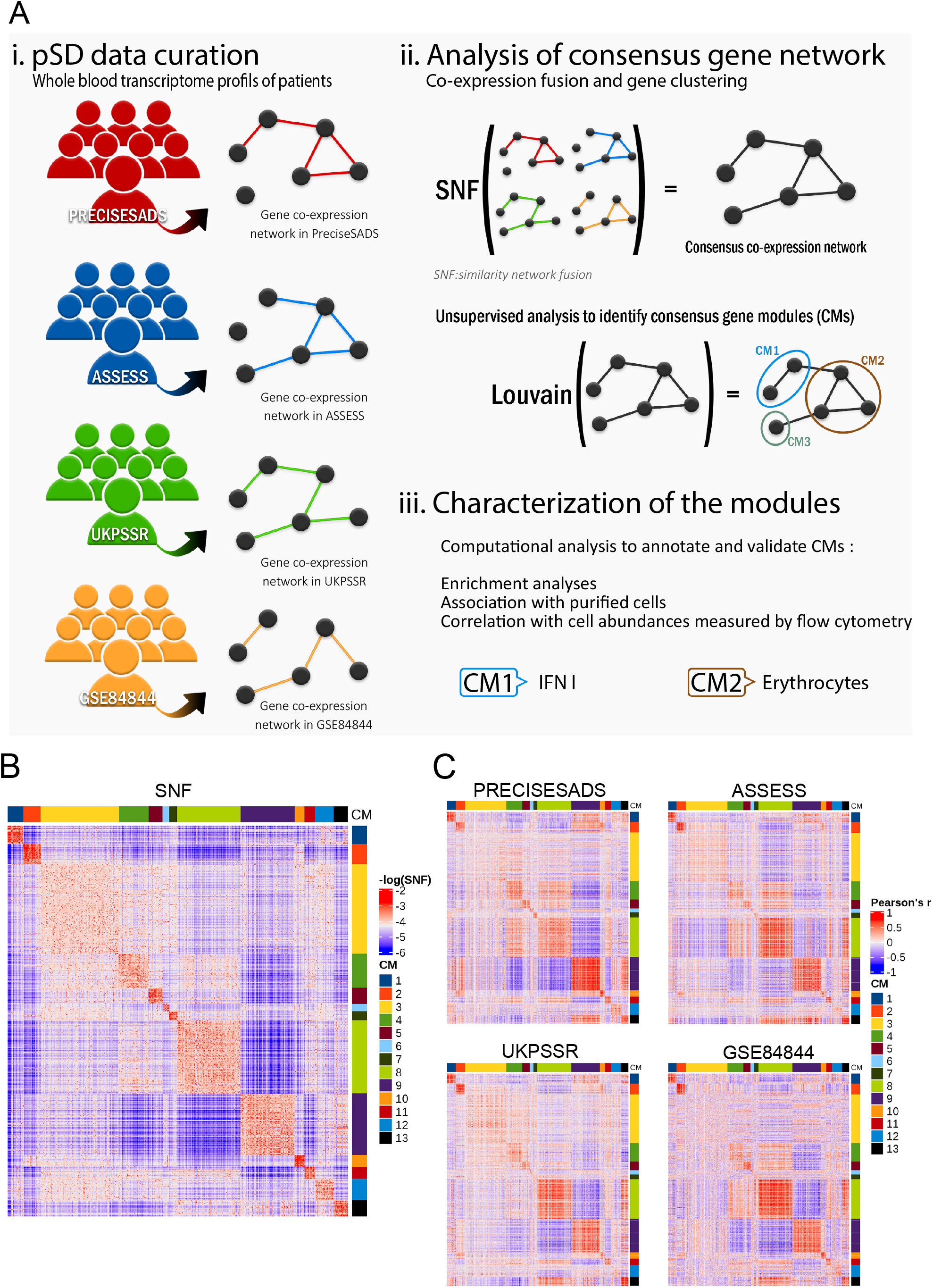
A) Schematic summary of the work. pSD = primary Sjögren Disease B) Heatmap of the consensus pairwise gene affinity computed by Similarity Network Fusion (SNF). Side annotations represent gene modules. C) Heatmaps of Pearson’s correlation matrices of the four input datasets, with genes grouped by their consensus gene modules.

### Biological interpretation of the CMs

The 13 CMs represent the main axes of heterogeneity of the blood transcriptome across pSD patients and can therefore facilitate the interpretation of high dimensional transcriptomic data by summarizing it using 13 dimensions. In order to biologically interpret these 13 axes of variation, we annotated each of them as corresponding to cell types or biological functions by using gene set enrichment analyses using gene sets from the Gene Ontology[27] and Altman *et al*.[28] databases (**Figure 2A, 2B**), as well as their average expression in transcriptomic profiles of sorted blood cell subsets[29] (**Figure 2C**).

**Fig. 2.**
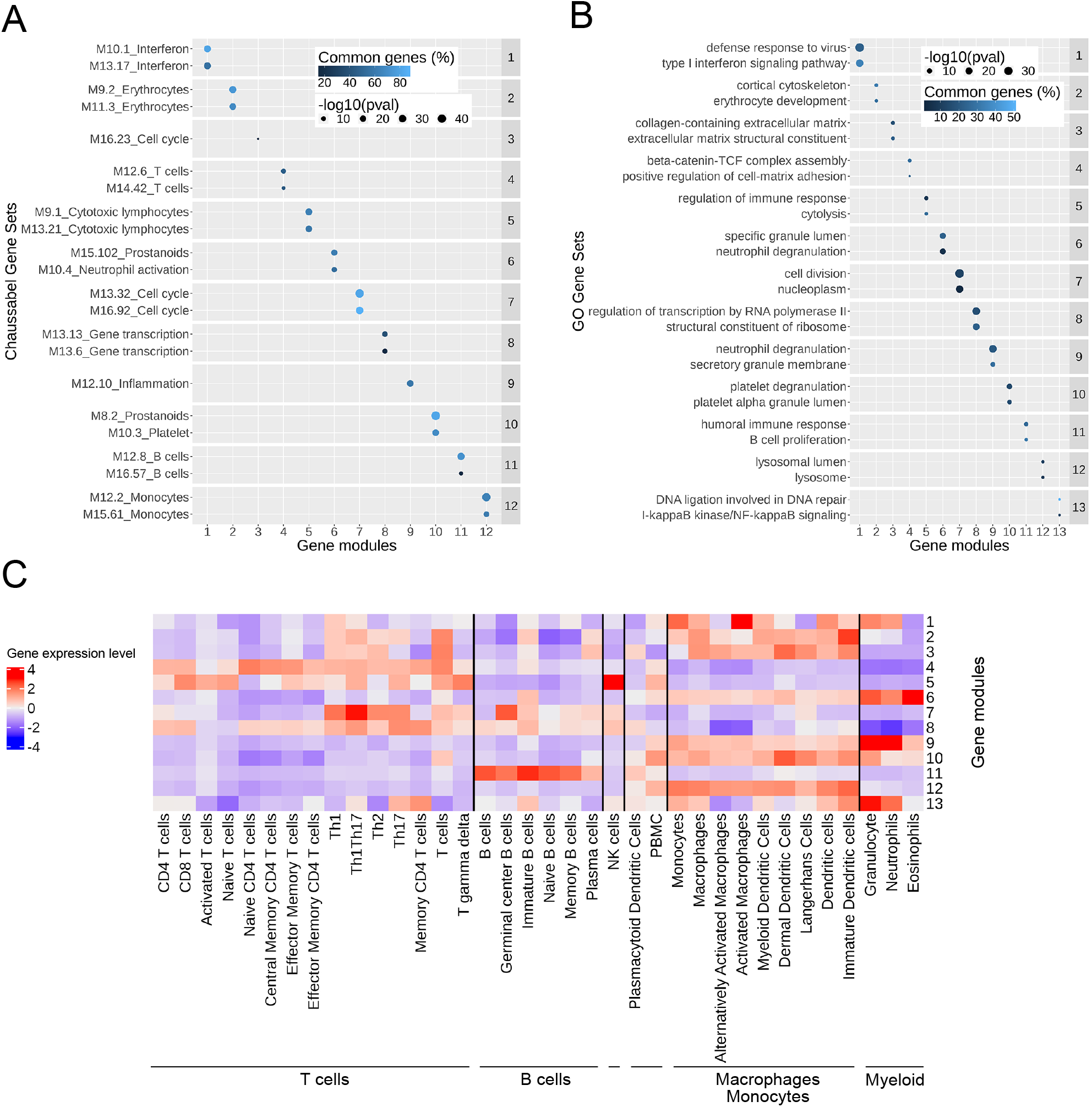
A) For each module, the two most significantly-enriched pathways in the Chaussabel database[**?**]. B) Most significantly-enriched pathways in the GO database[**?**] C) Average expression of modules in transcriptomes of purified cells

CM1 was enriched in Interferon related as well as response to viruses pathways, and we interpreted it as representing type 1 IFN signaling. CM7 was enriched in cell cycle-related genes, and we interpreted it as a transcriptomic signature of mitosis within blood cells.

Out of the 11 other modules, 9 represent different cell types. We found four modules corresponding to lymphoid cells: CM4, CM5 and CM11 were respectively enriched in pathways associated with T cells, NK cells and B cells functions **(Figure 2A, 2B)** and that were overexpressed in the transcriptome of the corresponding purified cell types **(Figure 2C)**. CM8 was enriched in genes associated with gene transcription and overexpressed across the transcriptomes of purified lymphocytes (T, B and NK cells) and therefore represents a shared gene transcription signature across all lymphocytes **(Figure 2C)**. We found six modules (CM2, CM6, CM9, CM10, CM12, CM13) representing myeloid cell subsets. CM2 was enriched in erythrocytes-annotated gene sets and CM10 in platelets-annotated gene sets. Module CM6 was overexpressed in the transcriptome of eosinophils. CM9 and CM13 were enriched in inflammation and neutrophil-related gene sets and overexpressed in the transcriptome of purified granulocytes and neutrophils. CM13 was in addition enriched in genes from the I-*κ*B kinase/NF-*κ*B signaling pathway, an inflammatory transcription factor expressed by neutrophils[30]. Finally, CM12 was enriched in gene sets related to monocytes and overexpressed in the transcriptome of cells derived from monocytes.

Among the 13 CMs, CM3, which contains the highest number of genes (n=1247), was the least co-expressed, had the lowest absolute expression levels **(Supplementary Figure 2)** module and showed inconsistent characterization results **(Figure 2A, 2B)**. We therefore did not take it into consideration for further analysis. In summary, we interpreted CM1 as type 1 interferon (IFN) activation, CM2 as representing the frequency of erythrocytes within the blood, CM3 as residual variance, CM4, CM5, CM6 as the frequencies of respectively T cells, NK cells and Eosinophils, CM7 as a signature of cell proliferation, CM8, CM10, CM11 and CM12 as the frequencies of respectively lymphocytes, platelets, B cells and monocytes, and CM9 and CM13 as representing neutrophils.

### Validation of the biological interpretations of the CMs

To confirm the biological interpretations of the CMs representing cell types, we compared their average expressions (**Material and Methods**) to the corresponding cellular frequencies measured by flow cytometry in matching samples whenever available (**Figure 3A**). For functional modules, we compared them to previously-published gene signatures (**Figure 3B**) or cytokines concentrations (**Figure 3C**).

**Fig. 3.**
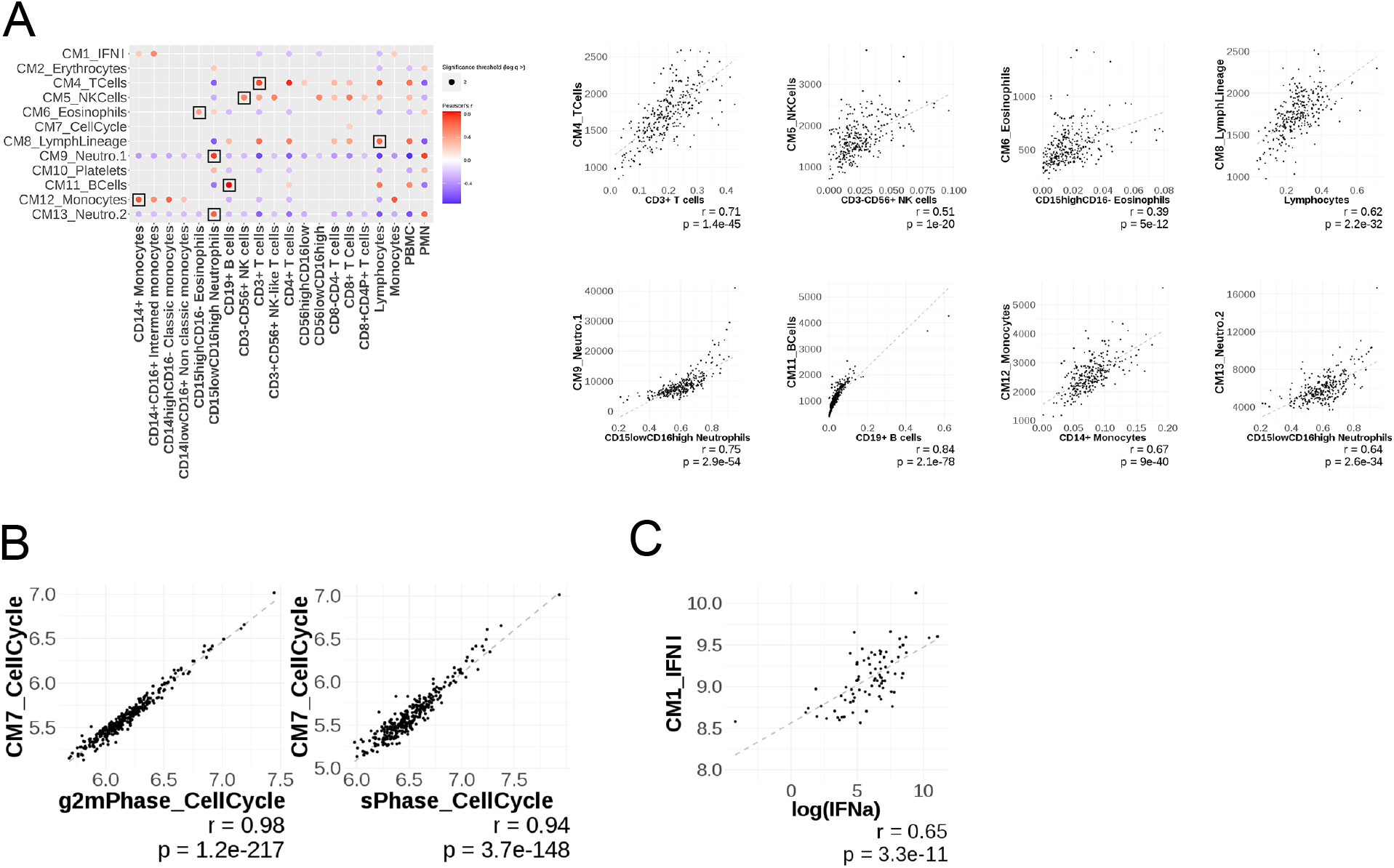
A) Significant Pearson’s correlations between the average expression of the CMs and cell types abundances measured by flow cytometry. Scatter plots of average CMs expression and matching cellular frequencies. B) Scatter plots illustrating the average expression of CM7 versus averages of cell cycle signatures C) Scatter plot of the average expression of CM1 IFN-*α* and dosage of IFN-*α*

For all the cellular modules for which we had matching cytometry data, we observed a high and significant correlation of the average module expression with the frequency among live single cells measured by flow cytometry (**Figure 3A**). More precisely, we observed correlation coefficients of 0.71 between the CM4 module and the frequency of T cells, of 0.51 between the CM5 modules and NK cells, of 0.39 between CM6 and eosinophils, 0.75 (respectively 0.64) between CM9 (respectively CM13) and neutrophils, 0.84 between CM11 and B cells, 0.67 between CM12 and monocytes, and 0.62 between CM8 and lymphocytes (all p-values *<* 2 *×* 10^−12^).

For functional modules, we observed a strong correlation (Pearson’s *r >* 0.94) of the CM7 with genes signatures corresponding to phases of the mitotic cycle identified with single cell RNA-sequencing data[31]. The other functional module CM1 was highly correlated with the concentration of type 1 IFN (measured by SIMOA) in the blood (*r* = 0.65, *p* = 3.3 *×* 10^−11^) (**Figure 3C**). Collectively, these analyses confirm the interpretation of the CMs derived from gene set enrichment analyses.

### The consensus gene modules identify consistency and heterogeneity across pSD patient stratifications

Three studies have proposed pSD patients stratifications according to molecular and clinical features of the disease[14–16]. Two methods were based on blood transcriptomic profiles of pSD patients on two distinct cohorts[15, 16]. Both studies identified four clusters of patients hereafter referred to as S1, S2, S3 and S4 (respectively T1, T2, T3 and T4) for the Soret (respectively Trutschel) classification. These stratifications were established using unsupervised clustering methods. Algorithmic classifiers to stratify new pSD cohorts according to these classification systems are however currently lacking, and no direct comparison has been performed so far.

Briefly, from Soret *et al*., cluster S1 exhibited high levels of interferon (IFN) activity and an increased frequency of B lymphocytes in the blood. Cluster S2 showed a similar expression profile to that of healthy volunteers. Cluster S3 displayed a high IFN signature, along with a more prominent involvement of B cell components compared to other clusters, including an increased frequency of B cells in the blood. Lastly, cluster C4 was characterized by an inflammatory signature driven by monocytes and neutrophils. Confirming the findings of *et al*.[15], our analysis confirmed the defining characteristics of these patient clusters. We consistently observed an upregulation of the Interferon module CM1 in S1 patients, the Neutrophils module CM9 in S4 patients, and the B cell module CM11 in S3 patients (Figure 4A). Our analysis further revealed that S3 is defined by a high abundance of lymphocytes (B, T, and NK cells represented by the CM11, CM4, and CM5 modules, respectively) associated with cell proliferation (CM7). Cluster S4 is characterized by a high abundance of platelets (CM10), erythrocytes (CM2), and neutrophils (CM9 and CM13). S1 is distinguished by high activation of type 1 IFN (CM1), while S2, described as normal-like by Soret *et al*., has fewer monocytes (CM12) and more T cells (CM4) compared to the cohort’s averages.

**Fig. 4.**
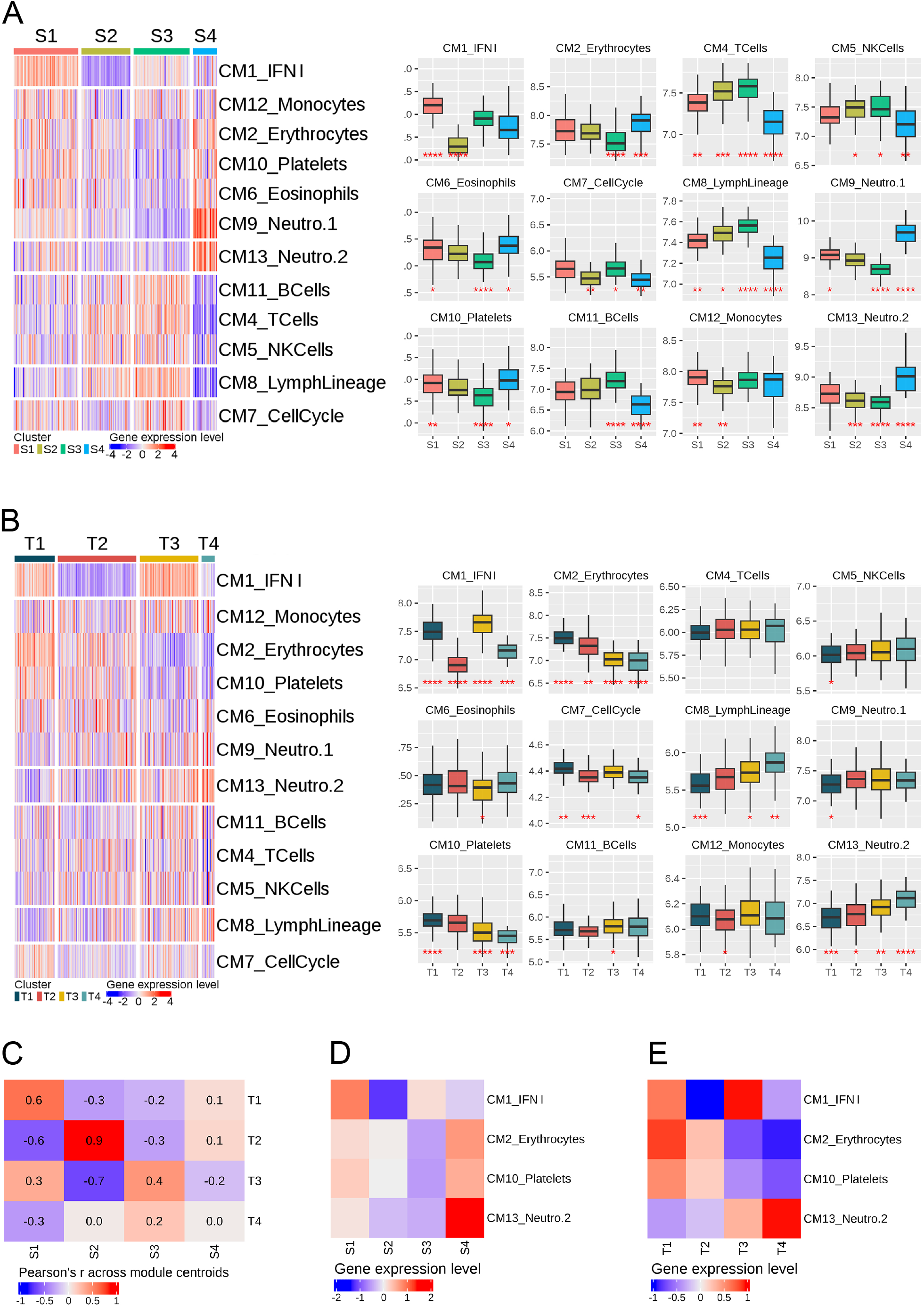
CMs scores across patient subgroups of A) the Soret classification B) the Trutschel classification and ANOVA tests for each clusters. Average expression of the CM1 IFN-*α*, CM2 Erythrocytes, CM10 Platelets and CM13 Neutrophils.2 CMs in the C) Soret classification and D) Trutschel classification. E) Correlation across cluster centroids of the two stratification systems.

In a separate study by Trutschel *et al*., four patient clusters were also identified. These clusters were based on two modules: IFN-stimulated genes (ISGs) and the erythroid module (ERM). Cluster T1 showed high expression of both these modules, while cluster T2 had low ISG expression but high ERM expression. Cluster T3 had high ISG expression and low ERM expression, and cluster T4 had low expression in both ISGs and ERM. We observed a high interferon signature (CM1) in clusters T1 and T3, with cluster T1 exhibiting a higher platelet presence compared to cluster T3 (Figure 4B). Cluster T2 had a lower abundance of monocytes (CM12), while cluster T4 had a high neutrophil signature (CM13). Cluster T1 had a high presence of erythrocytes, cluster T3 had fewer eosinophils (CM6), and clusters T3 and T4 had a higher abundance of lymphocytes (CM8).

To formally study the correspondence between the Soret and Trutschel classification systems, we computed Pearson correlation coefficients across centroids computed on mean-centered and unit variance-scaled module expression scores. This comparison highlighted a very high concordance between cluster S2 and T2 (r = 0.9), good concordance between clusters S1 and T1 (r = 0.6), moderate across clusters S3 and T3 (r = 0.4), and poor concordance across clusters S4 and T4 (r = 0) (**Figure 4E**). This analysis shows that there is a substantial overlap between the two classification systems, especially in the identification of T2 patients.

It therefore appears that cluster S1 of the Soret classification corresponds to cluster T1 of the Trutschel classification, marked by high type 1 IFN signaling (CM1) **(Figure 4C, 4D)**. Cluster S3 matches cluster T3, as identified by high type 1 IFN signaling (CM1) in the context of a lower abundance of platelets (CM10) and erythrocytes (CM2). Cluster S2 matches cluster T2, with the lowest type 1 IFN signature (CM1). Cluster S4 in resembles cluster T4, as both have the highest expression of the Neutrophil activation module (CM13), although other modules such as platelets (CM10) and erythrocytes (CM2) had discordant expression levels across the two patient classification systems. In general, there were no differences in the lymphoid modules (CM4, CM5 and CM11) across Trutschel clusters.

Tarn et al. propose a stratification model based on patient-reported symptoms and identified four clusters of patients: Low symptom burden (LSB), high symptom burden (HSB), dryness dominant with fatigue (DDF), and pain dominant with fatigue (PDF). We were unable to see any significant difference in the level of expression of any CM across the four subgroups of patients **(Supplementary Figure 3)**. Consistently, we observed -in the PreciseSADS and ASSESS cohorts-weak correlations of the CMs expression scores with the ESSDAI[32] and ESSPRI[33] disease activity scores **(Supplementary Figure 4)**. We however noted that unlike other components of the ESSDAI and ESSPRI disease activity scores, the presence of autoantibodies (anti-SSA, anti-SSB, PFLC, IgG) was positively-associated with the CM1 module representing type 1 IFN **(Supplementary Figure 5)**. These observations suggest that among pSD clinical manifestations, the presence of autoantibodies is the most associated with a specific blood transcriptomic profile.

### CM8 and CM2 are associated with response to hydroxychloroquine and leflunomide combination

Many clinical trials for Sjögren’s patients have shown poor results especially for response to treatment[34–37] but, negative clinical trials can still provide valuable information about the efficacy of a particular treatment and can help guide future research. However, positive trials provide a unique opportunity to compare responder and non-responder patients’ characteristics. Within the IMI2 NECESSITY, data from both positive and negative clinical trials are available for exploratory retrospective analyses. RepurpSS-1[38] is a placebo-controlled, double-blinded, phase 2A randomized clinical trial that evaluated the combination therapy of hydroxychloroquine and leflunomide and is one of the first positive clinical trials in pSD.

Firstly, we validated the co-expression of the genes within each CM on this cohort independent of those used for the identification of the modules, highlighting the reproducibility and generalizability of the CMs to independent pSD blood transcriptomic datasets **(Supplementary Figure 7)**.

Secondly, we looked at the evolution of the expression of each module between treatment initiation and completion. We observed that lefluonomide-hydroychloroquine combination led to a decrease in the expression of CMs representing T cells, platelets and B cells, and an increase expression of the CMs representing monocytes and neutrophils, thus suggesting that this treatment combination favored the number of myeloid immune cells over lymphoid immune cells in the blood (**Figure 5A**). While treatments received by patients before blood transcriptomic profiling were more heterogeneous in the PreciseSADS cohort, we consistently observed an influence of the type of treatment received on the expression level of the CMs (**Supplementary Figure 6**).

**Fig. 5.**
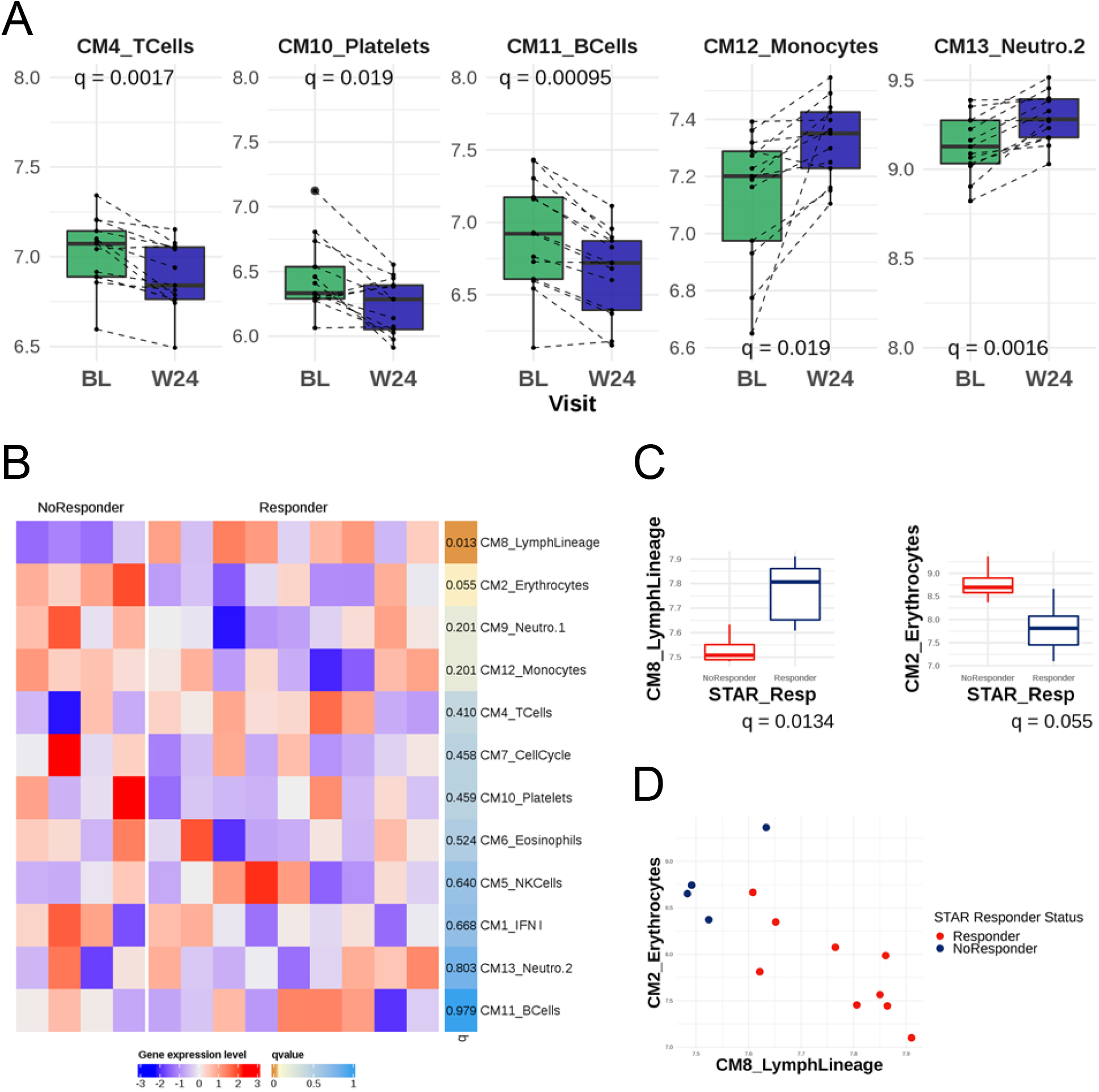
A) Boxplots illustrating the evolution of the modules significantly differentially-expressed at baseline (BL) versus Week 24 for treated patients B) Heatmap of average gene expression of the CMs. Patients are split by their responder status according to the STAR clinical endpoint C) Avegage expression of CM8 and CM2 at baseline in responders versus non-responders D) Dotplot of average expression of the CM8 and CM2 modules, colored by response statuses.

Finally, we examined whether the heterogeneity of the patients encompassed in the modules could help identify responders in the RepurpSS-1 trial before treatment initiation. To do so, we focused on the recently developed STAR clinical endpoint[39]. The CM8 Lymphoid Lineage module was significantly overexpressed in responders before treatment initiation (q = 0.013) (**Figure 5B, 5C, Supplementary Figure 8)**. Conversely, a trend for higher expression in non-responders of the CM2 module representing erythrocytes was also found (q = 0.055). By combining CM2 and CM8, we were able to perfectly separate responders and non-responders in this clinical trial (**Figure 5D**). These analyses suggest that these cell populations could represent biomarkers predictive of therapeutic efficacy of this treatment combination.

## Discussion

Primary Sjögren’s disease (pSD) is a debilitating and clinically heterogeneous disease with no well-established causal mechanism, nor approved targeted therapy. There is therefore an urgent need to identify biomarkers able to inform treatment selection as well as to stratify patients in clinical trials in the context of personalized medicine. High throughput transcriptomic profiling is an appealing technology for biomarker discovery as it allows the interrogation of tens of thousands of genes for differential expression across groups of patients, such as responders and non-responders to a drug in a clinical trial. The interpretation of transcriptomic profiles is however difficult, as groups of differentially expressed genes may represent dysregulation of functional pathways or changes in the cellular composition of samples, or both. In addition, the very high dimensionality of whole transcriptome assays makes difficult distinguishing true and replicable biological signal from noise.

To overcome these difficulties in the interpretation of the transcriptome in the context of pSD, we jointly analyzed four independent transcriptomic datasets profiling whole blood samples from pSD patients. We used clustering methods to identify the main axes of variation across these four datasets. As clustering algorithms are sensitive to noise, we implemented a method to perform a gene clustering analysis on a joint representation of the pairwise gene correlations matrix across the four datasets, rather than on each dataset separately. To do so, we recast the four observed matrices of pairwise gene correlations as graphs and used the SNF[25] algorithm to obtain a consensus graph representation of the gene correlation network across the four cohorts, on which we applied the Louvain graph clustering algorithm. We importantly showed that the gene modules we identified are reproducible across the four cohorts on which they were discovered **(Figure 1C)** as well as on an independent cohort (**Supplementary Figure 7**). These modules therefore represent the main biological features contained in the transcriptomic profile of the whole blood in pSD patients, therefore facilitating its interpretation for translational research.

In order to make the CMs more biological meaningful, we interpreted them using distinct public databases of pathways and blood cells transcriptomes[29]. This allowed us to identify both functional modules (interferon signaling or cell proliferation) or modules reflecting the cellular composition of the patients’ blood. Importantly, we observed highly significant correlations between the expression of the gene modules and corresponding cellular frequencies or cytokine levels, thus validating these computationally derived biological interpretations. In the recent years, so called transcriptomic deconvolution methods have been proposed in order to infer cellular proportions from transcriptomic measurements[40]. Most of these methods rely on a reference averaged transcriptomic profiles of cell types, usually derived from purified cells from the blood of healthy donors and use genes that are discriminative across cell populations in a given context, such as cancer[29]. In contrast, our approach is driven by the observed variations in the blood of pSD patients across multiple cohorts, ensuring that the gene signatures of the identified cell types are valid in this context. In addition, this data driven approach allowed us to define gene modules indicative of rare cell populations such as eosinophils or signatures of non-immune cell types such as erythrocytes or platelets which are not typically quantified by deconvolution algorithms[41]. Moreover, we found functional modules (CM1 type 1 IFN and CM7 Cell Cycle) that do not correspond to variations in the frequencies of blood cell types. The consensus gene modules described herein therefore could help understanding the complex pathophysiology of pSD as they represent biologically meaningful, reproducible, and sensitive sources of heterogeneity in the blood transcriptome of pSD patients.

The gene modules that we identified can serve as a building block for translational research in pSD, by providing a concise list of potential biomarkers provided by whole blood transcriptomic profiling. Multiple independent studies have recently focused on the stratification of the disease into discrete patient subgroups, based on whole blood transcriptomic profiles[15, 16] or clinical characteristics[14]. These classifications systems may become relevant in future clinical trials, as new treatments may benefit only to a restricted subset of patients. Our approach complements these classifications by highlighting the functional and cellular composition differences across patient subgroups, as well as highlighting the consensus and differences across classification systems. Our analyses notably suggest that the patient subgroups in published transcriptomic-based patient stratification systems can be distinguished based on the measurement of three variables: the frequency of neutrophils in the peripheral blood, the concentration of type 1 IFN, as well as the frequency of either erythrocytes or platelets within the blood (**Figure 4C, 4D**). These observed differences across patient subgroups may provide clinically actionable biomarkers for disease stratification in settings where whole blood transcriptomic profiling is impractical. Indeed, these key features of pSD drive disease heterogeneity and altogether may be useful predictors of response.

Some medications are designed to target specific genes or proteins, altering their activities and ultimately leading to changes in cellular behavior. Understanding the complex relationship between medications and gene expression is an important area of research that includes Drug Repurposing computational activities and may eventually lead to the definition of more effective treatment strategies for a wide range of diseases and conditions. Our analyses showed that the CMs can be used to understand the effect of drugs on the composition and functional orientation of the peripheral blood **(Figure 5, Supplementary Figure 7)**. We also confirmed, in two independent cohorts, the correlation between the presence of anti-SSA and anti-SSB autoantibodies and the level of type 1 IFN in the peripheral blood. The pathogenic role of the IFN pathway has been extensively described: type I IFN signature is correlated with the development of systemic extra-glandular manifestations, and a substantial production of autoantibodies and inflammatory cytokines[42]. Moreover, in the context of systemic autoimmune manifestations, pSD patients may present with hematologic abnormalities including anaemia, leukopenia (mainly neutropenia or lymphopenia), and thrombocytopenia[43, 44]. These three components are indeed evaluated in the haematological domain of the ESSDAI scale. As these patient characteristics are recapitulated by our CMs, whole blood transcriptomic profiling thus appears informative in the context of pSD translational research.

The CMs we identified indeed provide a succinct list of candidate blood-based biomarkers that recapitulate whole transcriptome profiles in a biologically interpretable manner. These modules can therefore be examined in exploratory and clinical research for their potential association with the response to a treatment or to study drug mechanism of action. We exemplified this idea by retrospectively analyzing data from the RepurpSS-1 phase IIa clinical trial[38] which evaluated a combination of leflunomide and hydroxychloroquine for the treatment of pSD. Longitudinal whole blood transcriptomic profiling allowed us to show that this combination led to a decreased expression of CMs corresponding to T cells, platelets and B cells, and an increase in modules representing monocytes and neutrophils. Our results therefore show that this combination of treatments influence the cellular composition of the peripheral blood in pSD patients.

Importantly, we investigated the relationship between each CM expression levels before treatment initiation and the observed clinical response upon completion of the clinical trial. Our results show that responders to this treatment combination featured higher expression of the module representing lymphocytes and a trend for lower expression of the module representing erythrocytes. These observations are consistent with the mechanism of action of leflunomide, an immunomodulatory drug known to inhibit de novo synthesis of pyrimidine, preventing lymphocytes from expanding in inflammatory context[45]. While the mechanism of hydroxychloroquine is less clear considering its initial use as an antimalarial drug, this molecule has widely been used in rheumatic autoimmune diseases such as systemic lupus erythematosus[46]. Studies have shown that hydroxychloroquine can contribute to regulate inflammation by blocking Toll-like receptors (TLR) leading to type I IFN pathway inhibition[47]. Hydroxychloroquine has also demonstrated inhibitory effect on platelet activation[48], in accordance with modulations seen on CM relating to platelets in the RepurpSS-1 clinical trial. Our results suggest that clinical efficacy for this treatment combination may be restricted to patients with high lymphoid frequency and low erythrocytes frequency, thus providing new hypotheses guiding the treatment strategy of pSD patients and the design of future clinical trials.

Our work is therefore expected to facilitate translational and clinical research on primary Sjögren’s disease by presenting a set of reproducible and annotated gene modules that capture the major variations in the blood transcriptome of patients, which will open up the path for identifying biomarkers in clinical trials for this disease that is still poorly managed.

## Data Availability

All data produced in the present study are available upon reasonable request to the authors

## Author contributions

Conceptualization: C.B., M.G., E.Be, E.Bi, L.L.

Methodology: C.B., M.G., E.Be, E.Bi, A.H., B.C.

Validation: D.T, P.S, E.D.

Formal analysis: C.B, A.H, B.C., A.B, E.Be

Writing – Original Draft: C.B., E.Be, E.Bi, L.L.

Writing – Review and Editing: P.S., D.T., A.H., B.C, C.L., A.A., S.H., P.M., E.D, A.B, M.A.R, W.F.N, J.R, J-E.G., B.S., X.M, M.G.

Resources: J-E.G, M.E.A.R, W.F.N, J.R.

Supervision: M.G., E.Be, E.Bi, L.L.

We, the authors of this manuscript, confirm that we have collectively agreed to submit this work for publication. We have read and approved the final draft and take full responsibility for its content, including the accuracy of the data presented. We have also ensured that the statistical analysis, where applicable, was conducted appropriately and accurately. As authors, we are committed to upholding the highest standards of scientific integrity and ethical conduct, and we affirm that this work represents our best efforts to contribute to the advancement of knowledge in our field.

## Figures

**Figure 1 A)** Schematic summary of the work. pSD = primary Sjögren Disease **B)** Heatmap of the consensus pairwise gene affinity computed by Similarity Network Fusion (SNF). Side annotations represent gene modules.**C)** Heatmaps of Pearson’s correlation matrices of the four input datasets, with genes grouped by their consensus gene modules.

Figure 2. A) For each module, the two most significantly-enriched pathways in the Chaussabel database[28]. B) Most significantly-enriched pathways in the GO database[27] C) Average expression of modules in transcriptomes of purified cells

Figure 3. A) Significant Pearson’s correlations between the average expression of the CMs and cell types abundances measured by flow cytometry. Scatter plots of average CMs expression and matching cellular frequencies. B) Scatter plots illustrating the average expression of CM7 versus averages of cell cycle signatures C) Scatter plot of the average expression of CM1 type 1 IFN and dosage of type 1 IFN

Figure 4. CMs scores across patient subgroups of A) the Soret classification B) the Trutschel classification. Average expression of the CM1 type 1 IFN, CM2 Erythrocytes, CM10 Platelets and CM13 Neutrophils.2 CMs in the C) Soret classification and D) Trutschel classification. E) Correlation across cluster centroids of the two stratification systems.

Figure 5. A) Boxplots illustrating the evolution of the modules significantly differentially-expressed at baseline (BL) versus Week 24 for treated patients B) Heatmap of baseline average gene expression of the CMs. Patients are split by their responder status according to the STAR clinical endpoint. Right side annotations indicate FDR corrected p-value (qvalue) C) Avegage expression of CM8 and CM2 at baseline in responders versus non-responders D) Dotplot of average expression of the CM8 and CM2 modules, colored by response statuses.

## Material and Methods

### Data collection

Gene expression and associated clinical and biological data was obtained through tranSMART, the NECESSITY consortium data sharing platform for the ASSESS (Assessment of Systemic complications and Evolution in Sjögren’s Syndrome) cohort[22], PRECISESADS[12] and UKPSSR[23] cohort. Data from the fourth cohort was downloaded from the Gene Expression Omnibus repository, under the accession number GSE84844[24].

### Transcriptomic data pre-processing

The UKPSSR RNA-seq count data was transformed as in[14]. RNA-seq data from the PreciseSADS cohort was normalized as in Soret *et al*.[15]. The ASSESS Affymetrix Clariom S microarray data were normalized as in[16].

The GSE84844 Affymetrix Human Genome U133 Plus 2.0 Array data was pre-treated by filtering out probesets indistinguishable from background noise. For that purpose, we modeled probesets expression after applying a *log*2(*x* + 1) transformation by a two component Gaussian mixture model[**dempster•maximum•1977**] with the first peak corresponding to unexpressed genes, and the second peak to expressed genes. We retrieved the parameters of the mixture distribution using the function *normalmixEM* from the *mixtools* R package. The 0.95^*t*^*h* quantile of the first component of the distribution was used as a threshold. Probesets whose expression were below that threshold in more than 95% of the samples were removed. Finally, the fRMA function from the fRMA R Package[**McCall•Bolstad•Irizarry•1970**] was used to normalize probesets intensities across samples.

Finally, to have comparable data sets, the intersection of the 80% most varying common genes across all the data sets was selected (5443 genes).

### Integrated affinity network

The construction of the integrated network involves two steps: First, gene affinity (*affi*) is computed independently on each data set as follow : for each pair of genes (*x, y*), we consider the affinity between *x* and *y* as *affi*_(*x,y*)_ = *exp*((1 − *cor*(*x, y*))*/σ*) where *cor* is the Pearson correlation coefficient and *σ* = 3, as suggested by Wang *et al*.[25]. The four networks are then merged into an integrated affinity network by using the Similarity Network Fusion (SNF) method[25], with 30 neighbours per gene and 20 iterations. The SNF algorithm produces a weighted fully connected graph with 5000^2^ = 2.5 *×* 10^6^ edges. Visual inspection of the distribution of the weights showed that their distribution was bimodal, with a largely preponderant low weight peak **[Supplementary Figure 1]**. To convert the fully connetected output of the SNF algorithm to a sparse graph, we removed edges below the 0.9775*th* quantile of the weights distribution **(Supplementary Figure 1)**.

### Consensus modules identification

Consensus gene modules were identified by applying the Louvain clustering algorithm[26] on the fused and truncated graph of pairwise gene affinities. This method is based on a modularity optimization algorithm that aims to partition genes into communities with high within-group affinity and low between-group affinity. The modularity score of a community structure is calculated as the difference between the weighted proportion of intra-community edges and the expected weighted proportion of such edges if the edges were randomly distributed.

### Gene modules summarization

We used the mean expression the genes contained in a module to represent that module’s expression as performed in Becht et al[29].

### Gene set enrichment analysis

Enrichment analysis is performed by applying a Fisher-exact tests on the human blood-derived transcriptomic modules of Altman *et al*.[28] as well as the Gene Ontology database[27]. P-values were corrected using the Benjamini-Hochberg procedure to select pathways by controlling the false discovery rate at a 0.05 level.

### Mapping with purified and sorted immune cells

To identify modules representing the abundances of blood cell types, we used the GSE86362 dataset[29], which consists of 1936 gene expression profiles from immune cell populations, non-immune non-malignant cell populations and non-hematopoietic cancer cell lines. For consistency with our sample types, we only retained samples corresponding to blood cell populations (*n* = 1095).

### Correlation between CMs and cell type abundances measured by Flow Cytometry

On the PreciseSADS cohort, proportions of relevant cell types using flow cytometry custom marker panels were analyzed for samples where matched transcriptomic profiles and cytometry data were available. Correlations were performed between summarized CM expression levels and log-frequencies of the corresponding cell populations among live single cells, as previously described[29]. We corrected the p-values by Benjamini-Hochberg (BH) procedure by controlling the False Discovery Rate (FDR) at a 0.05 level.

### Correlation between CMs and cytokines

On the PreciseSADS cohort, relevant cytokines were measured as in[12]. A log transformation was applied on the concentrations. Finally, we computed correlations tests between the average expression of the CMs and the cytokines levels we corrected the p-value by controlling the FDR at a 0.05 level (BH procedure).

### Application to clinical trial

RepurpSS-1 (registered under trial number EudraCT, 2014–003140–12) was a phase II a placebo-controlled clinical trial testing a combination of Leflunomide and Hydroxychloroquine[38]. Gene expression and associated biological and clinical data for the RepurpSS-1 trial was obtained through the NECESSITY consortium. Transcriptomes of samples with a RIN *<* 6 or DV200 *>* 70 were excluded, resulting in the analysis of 16 patients. Pre-treatment and post-treatment (at week 24) CM expression levels were compared using paired t-tests with Benjamini-Hochberg correction. Responder status was determined based on the STAR clinical composite endpoint[39]. Patients with a STAR score of 5 or above were classified as responders. Difference in CM expression levels between responders and non-responders were assessed using univariate t-tests with BH FDR correction.

## Supplementary materials

**Table 1**. List of genes (SYMBOL) in each Concensus Modules (CMs)

**Supplementary Fig1**. Histogram showing the distribution of weights in the SNF matrix. The x-axis denotes the weight range (logged) and the y-axis represents the frequency of weights. A vertical red line indicates the discretization threshold corresponding to the 0.975^*th*^ quantile (for better visualization).

**Supplementary Fig2. A)**Average correlation of the 4 input datasets **B)** Average of average correlation matrices **C)** Average gene expression levels for each CM in cohorts profiled by RNA-sequencing

**Supplementary Fig3**. CMs scores across patient subgroups of the Tarn classification in UKPSSR cohort

**Supplementary Fig4**. Pearson’s correlation between average CMs expression and ESSDAI and ESSPRI scores in A) PRECISESADS and B) ASSESS cohorts

**Supplementary Fig5**. Pearson’s correlation between average CMs expression and autoantibodies levels in A) PRECISESADS and B) ASSESS cohorts

**Supplementary Fig6. A)**T-test between average CMs expression and treatment. q = corrected p-value **B)**CMs expression scores across patients stratified by treatments received. AM = Antimalarials, STD = Steroids, IS = Immunosupressors **C)**Significant diffrences observed in treated versus untreated patients.

**Supplementary Fig7**. Correlation matrix in REPURPSS-1 cohort, sorted by CMs.

**Supplementary Fig9**. Boxplots of average expression of the CMs at baseline versus after treatment splitting patients by treatment and placebo.

**Supplementary Fig8**. Boxplots of average expression of the CMs versus response status.

## References

1. Mariette, X. & Criswell, L. A. Primary Sjögren’s Syndrome. New England Journal of Medicine 378 (ed Solomon, C. G.) 931–939 (Mar. 2018).

2. Brito-Zerón, P. et al. Sjögren syndrome. Nature Reviews Disease Primers 2 (July 2016).

3. Parisis, D., Chivasso, C., Perret, J., Soyfoo, M. S. & Delporte, C. Current State of Knowledge on Primary Sjögren’s Syndrome, an Autoimmune Exocrinopathy. Journal of Clinical Medicine 9, 2299 (July 2020).

4. Solans-Laqué, R. et al. Risk, Predictors, and Clinical Characteristics of Lymphoma Development in Primary Sjögren’s Syndrome. Seminars in Arthritis and Rheumatism 41, 415–423 (Dec. 2011).

5. Nocturne, G., Pontarini, E., Bombardieri, M. & Mariette, X. Lymphomas complicating primary Sjögren’s syndrome: from autoimmunity to lymphoma. Rheumatology (Mar. 2019).

6. Narváez, J., Sánchez-Fernández, S. Á., Seoane-Mato, D., Dıaz-González, F. & Bustabad, S. Prevalence of Sjögren’s syndrome in the general adult population in Spain: estimating the proportion of undiagnosed cases. Scientific Reports 10 (June 2020).

7. Mavragani, C. P. & Moutsopoulos, H. M. The geoepidemiology of Sjögren’s syndrome. Autoimmunity Reviews 9, A305–A310 (Mar. 2010).

8. Anagnostopoulos, I. et al. The prevalence of rheumatic diseases in central Greece: a population survey. BMC Musculoskeletal Disorders 11 (May 2010).

9. Maldini, C. et al. Epidemiology of Primary Sjögren’s Syndrome in a French Multiracial/Multiethnic Area. Arthritis Care &amp Research 66, 454–463 (Feb. 2014).

10. Vivino, F. B. Sjogren’s syndrome: Clinical aspects. Clinical Immunology 182, 48–54 (Sept. 2017).

11. Qin, B. et al. Epidemiology of primary Sjögren’s syndrome: a systematic review and meta-analysis. Annals of the Rheumatic Diseases 74, 1983–1989 (June 2014).

12. Barturen, G., Beretta, L., Cervera, R., Vollenhoven, R. V. & Alarcón-Riquelme, M. E. Moving towards a molecular taxonomy of autoimmune rheumatic diseases. Nature Reviews Rheumatology 14, 75–93 (Jan. 2018).

13. Barturen, G. et al. Integrative Analysis Reveals a Molecular Stratification of Systemic Autoimmune Diseases. Arthritis &amp Rheumatology 73, 1073–1085 (Apr. 2021).

14. Tarn, J. R. et al. Symptom-based stratification of patients with primary Sjögren’s syndrome: multi-dimensional characterisation of international observational cohorts and reanalyses of randomised clinical trials. The Lancet Rheumatology 1, e85–e94 (Oct. 2019).

15. Soret, P. et al. A new molecular classification to drive precision treatment strategies in primary Sjögren’s syndrome. Nature Communications 12 (June 2021).

16. Trutschel, D. et al. Variability of Primary Sjögren’s Syndrome Is Driven by Interferon-α and Interferon-α Blood Levels Are Associated With the Class II HLA–DQ Locus. Arthritis &amp Rheumatology 74, 1991–2002 (Nov. 2022).

17. Fu, X., Liu, H., Huang, G. & Dai, S.-S. The emerging role of neutrophils in autoimmune-associated disorders: effector, predictor, and therapeutic targets. MedComm 2, 402–413 (July 2021).

18. Negrini, S. et al. Sjögren’s syndrome: a systemic autoimmune disease. Clinical and Experimental Medicine 22, 9–25 (June 2021).

19. Bombardieri, M. et al. One year in review 2020: pathogenesis of primary Sjögren’s syndrome. Clinical and experimental rheumatology 38 Suppl 126, 3–9. ISSN: 0392-856X (4 Jul-Aug 2020). ppublish.

20. Saraux, A., Pers, J.-O. & Devauchelle-Pensec, V. Treatment of primary Sjögren syndrome. Nature Reviews Rheumatology 12, 456–471 (July 2016).

21. Ritter, J., Chen, Y., Stefanski, A.-L. & Dörner, T. Current and future treatment in primary Sjögren’s syndrome – A still challenging development. Joint Bone Spine 89, 105406 (Nov. 2022).

22. Gottenberg, J.-E. et al./person-group>. Serum Levels of Beta2-Microglobulin and Free Light Chains of Immunoglobulins Are Associated with Systemic Disease Activity in Primary Sjögren’s Syndrome. Data at Enrollment in the Prospective ASSESS Cohort. PLoS ONE 8 (ed Re, V. D.) e59868 (May 2013).

23. Ng, W.-F., Bowman, S. J. & and, B. G. United Kingdom Primary Sjogren’s Syndrome Registry–a united effort to tackle an orphan rheumatic disease. Rheumatology 50, 32–39 (Aug. 2010).

24. Tasaki, S. et al. Multiomic disease signatures converge to cytotoxic CD8 T cells in primary Sjögren’s syndrome. Annals of the Rheumatic Diseases 76, 1458–1466 (May 2017).

25. Wang, B. et al. Similarity network fusion for aggregating data types on a genomic scale. Nature Methods 11, 333–337 (Jan. 2014).

26. Blondel, V. D., Guillaume, J.-L., Lambiotte, R. & Lefebvre, E. Fast unfolding of communities in large networks. Journal of Statistical Mechanics: Theory and Experiment 2008, P10008 (Oct. 2008).

27. Ashburner, M. et al. Gene Ontology: tool for the unification of biology. Nature Genetics 25, 25–29 (May 2000).

28. Altman, M. C. et al. Development of a fixed module repertoire for the analysis and interpretation of blood transcriptome data. Nature Communications 12 (July 2021).

29. Becht, E. et al. Estimating the population abundance of tissue-infiltrating immune and stromal cell populations using gene expression. Genome Biology 17 (Oct. 2016).

30. Castro-Alcaraz, S., Miskolci, V., Kalasapudi, B., Davidson, D. & Vancurova, I. NF-κB Regulation in Human Neutrophils by Nuclear IκBα: Correlation to Apoptosis. The Journal of Immunology 169, 3947–3953 (Oct. 2002).

31. Tirosh, I. et al. Dissecting the multicellular ecosystem of metastatic melanoma by single-cell RNA-seq. Science 352, 189–196 (Apr. 2016).

32. Seror, R. et al. EULAR Sjögren’s syndrome disease activity index: development of a consensus systemic disease activity index for primary Sjögren’s syndrome. Annals of the Rheumatic Diseases 69, 1103–1109 (June 2009).

33. Seror, R. et al. EULAR Sjögren’s Syndrome Patient Reported Index (ESSPRI): development of a consensus patient index for primary Sjögren’s syndrome. Annals of the Rheumatic Diseases 70, 968–972 (Feb. 2011).

34. Devauchelle-Pensec, V. et al. Treatment of Primary Sjögren Syndrome With Rituximab. Annals of Internal Medicine 160, 233–242 (Feb. 2014).

35. Bowman, S. J. et al. Randomized Controlled Trial of Rituximab and Cost-Effectiveness Analysis in Treating Fatigue and Oral Dryness in Primary Sjögren’s Syndrome. Arthritis &amp Rheumatology 69, 1440–1450 (June 2017).

36. Ship, J. A. et al. Treatment of Primary Sjogren’s Syndrome with Low-Dose Natural Human Interferon-alpha Administered by the Oral Mucosal Route: A Phase II Clinical Trial. Journal of Interferon &amp Cytokine Research 19, 943–951 (Aug. 1999).

37. Zandbelt, M. M. et al. Etanercept in the treatment of patients with primary Sjögren’s syndrome: a pilot study. The Journal of rheumatology 31, 96–101. ISSN: 0315-162X (1 Jan. 2004). ppublish.

38. Van der Heijden, E. H. M. et al. Leflunomide–hydroxychloroquine combination therapy in patients with primary Sjögren’s syndrome (RepurpSS-I): a placebo-controlled, double-blinded, randomised clinical trial. The Lancet Rheumatology 2, e260–e269 (May 2020).

39. Seror, R. et al. Development and preliminary validation of the Sjögren’s Tool for Assessing Response (STAR): a consensual composite score for assessing treatment effect in primary Sjögren’s syndrome. Annals of the Rheumatic Diseases 81, 979–989 (Apr. 2022).

40. Finotello, F. & Trajanoski, Z. Quantifying tumor-infiltrating immune cells from transcriptomics data. Cancer Immunology, Immunotherapy 67, 1031–1040 (Mar. 2018).

41. Sturm, G. et al. Comprehensive evaluation of transcriptome-based cell-type quantification methods for immuno-oncology. Bioinformatics 35, i436–i445 (July 2019).

42. Papa, N. D. et al. The Role of Interferons in the Pathogenesis of Sjögren’s Syndrome and Future Therapeutic Perspectives. Biomolecules 11, 251 (Feb. 2021).

43. Stergiou, I. E., Kapsogeorgou, E. E., Tzioufas, A. G., Voulgarelis, M. & Goules, A. V. Clinical Phenotype and Mechanisms of Leukopenia/Neutropenia in Patients with Primary Sjögren’s Syndrome. Mediterranean Journal of Rheumatology 33, 99 (2022).

44. Wen, W. et al. Clinical and serologic features of primary Sjögren’s syndrome concomitant with autoimmune hemolytic anemia: a large-scale cross-sectional study. Clinical Rheumatology 34, 1877–1884 (Oct. 2015).

45. Breedveld, F. C. Leflunomide: mode of action in the treatment of rheumatoid arthritis. Annals of the Rheumatic Diseases 59, 841–849 (Nov. 2000).

46. Shippey, E. A., Wagler, V. D. & Collamer, A. N. Hydroxychloroquine: An old drug with new relevance. Cleveland Clinic Journal of Medicine 85, 459–467 (June 2018).

47. Kužnik, A. et al. Mechanism of Endosomal TLR Inhibition by Antimalarial Drugs and Imidazoquinolines. The Journal of Immunology 186, 4794–4804 (Apr. 2011).

48. Erkan, D. et al. 14th International Congress on Antiphospholipid Antibodies Task Force Report on Antiphospholipid Syndrome Treatment Trends. Autoimmunity Reviews 13, 685–696 (June 2014).

