## Supplementary Figures for "Consensus gene modules strategy identifies candidate blood-based biomarkers for primary Sjögren’s disease"

### Supplementary materials

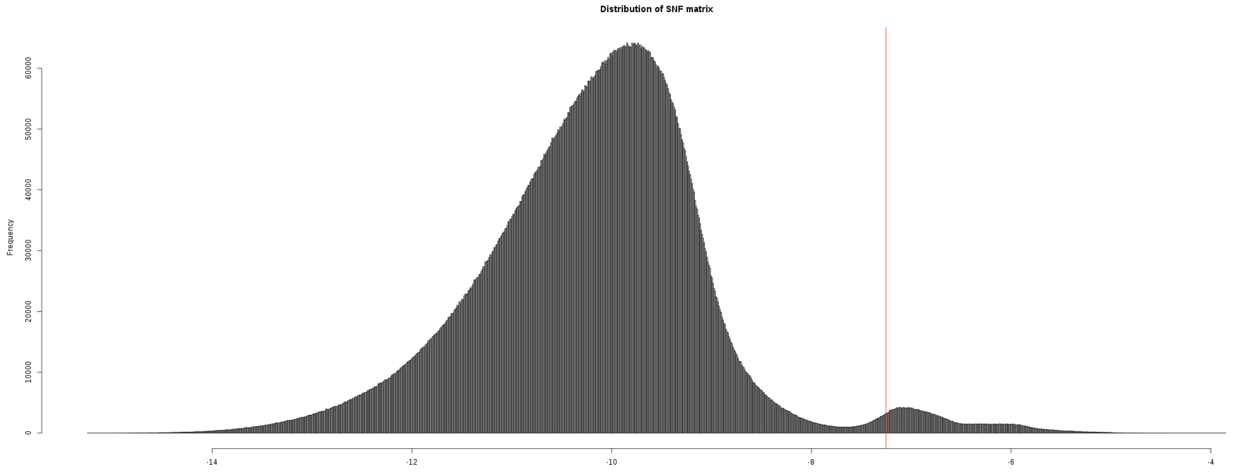


**Supplementary Fig1.** Histogram showing the distribution of weights in the SNF matrix. The x-axis denotes the weight range (logged) and the y-axis represents the frequency of weights. A vertical red line indicates the discretization threshold corresponding to the $0.975^{th}$ quantile (for better visualization).

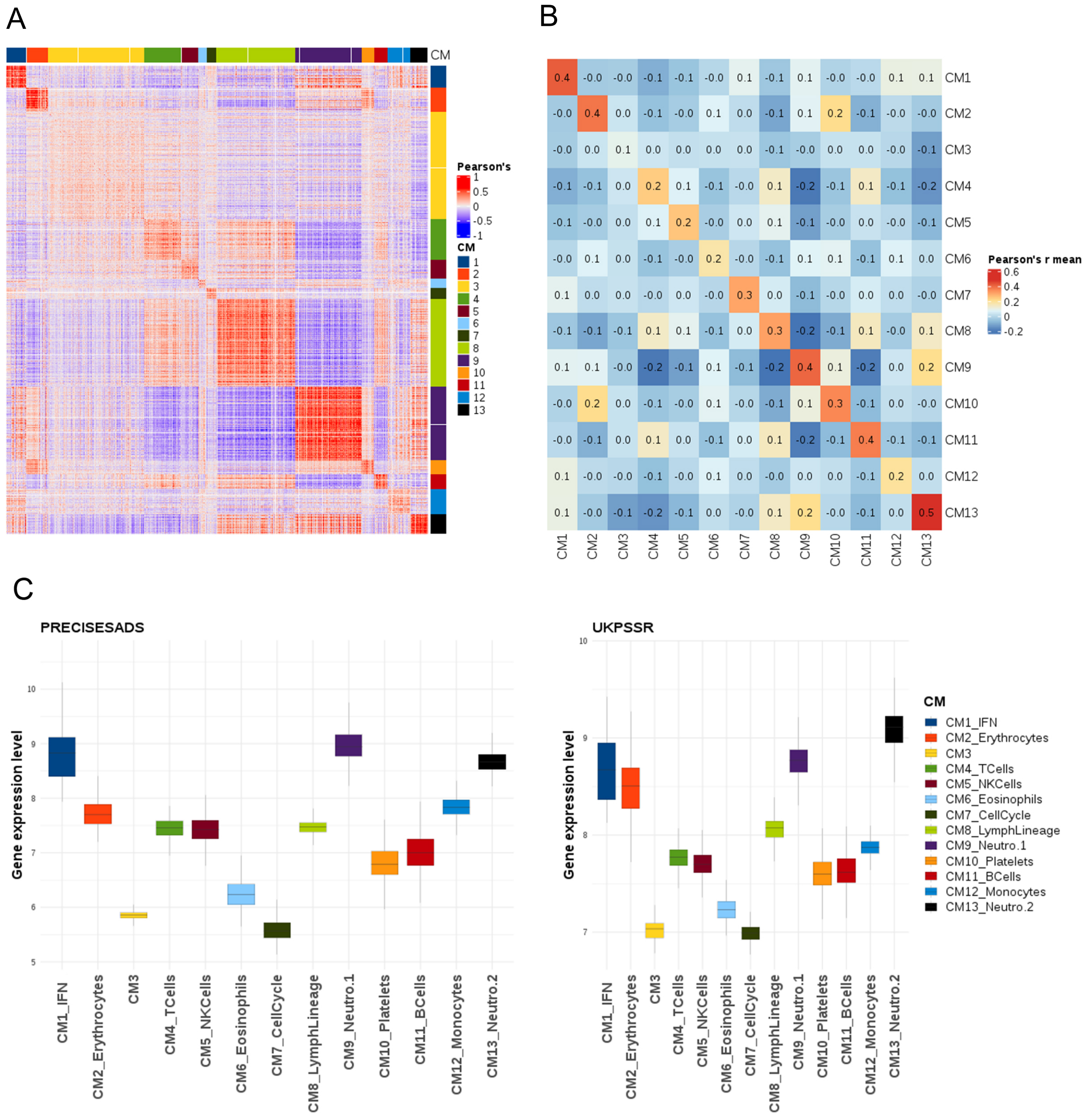


**Supplementary Fig2. A)**Average correlation of the 4 input datasets **B)** Average of average correlation matrices **C)** Average gene expression levels for each CM in cohorts profiled by RNA-sequencing

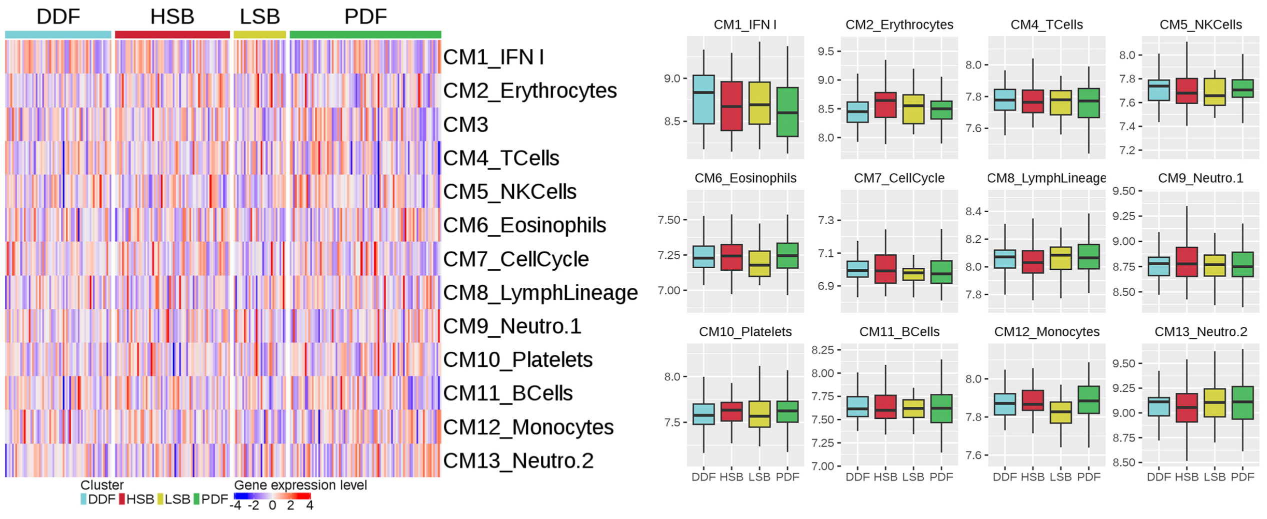


**Supplementary Fig3.** CMs scores across patient subgroups of the Tarn classification in UKPSSR cohort

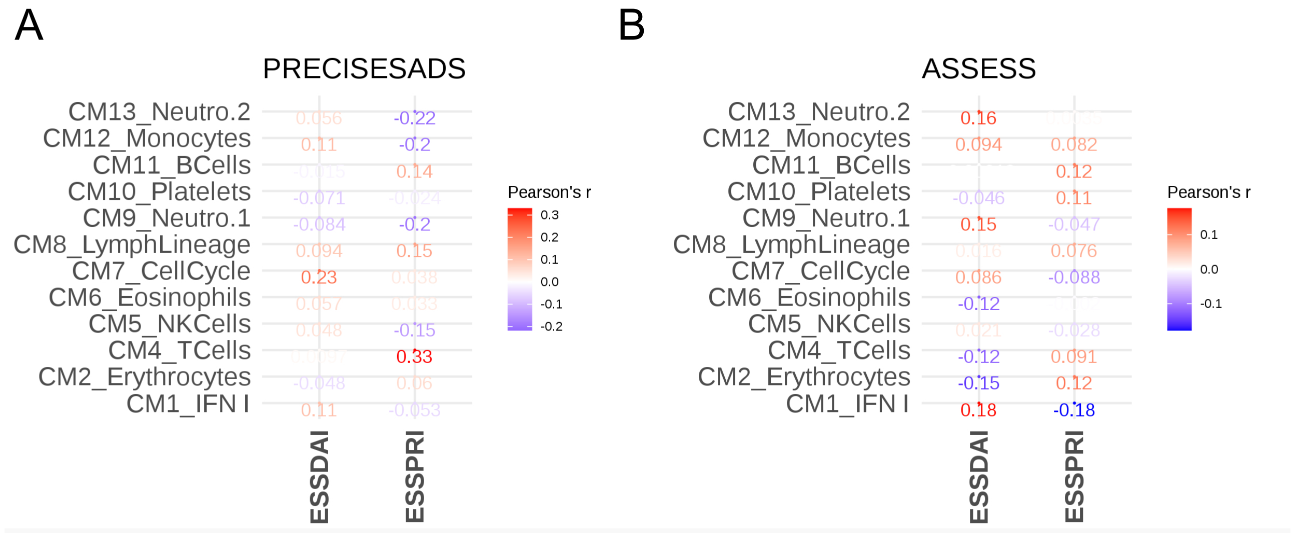


**Supplementary Fig4.** Pearson's correlation between average CMs expression and ESSDAI and ESSPRI scores in A) PRECISESADS and B) ASSESS cohorts

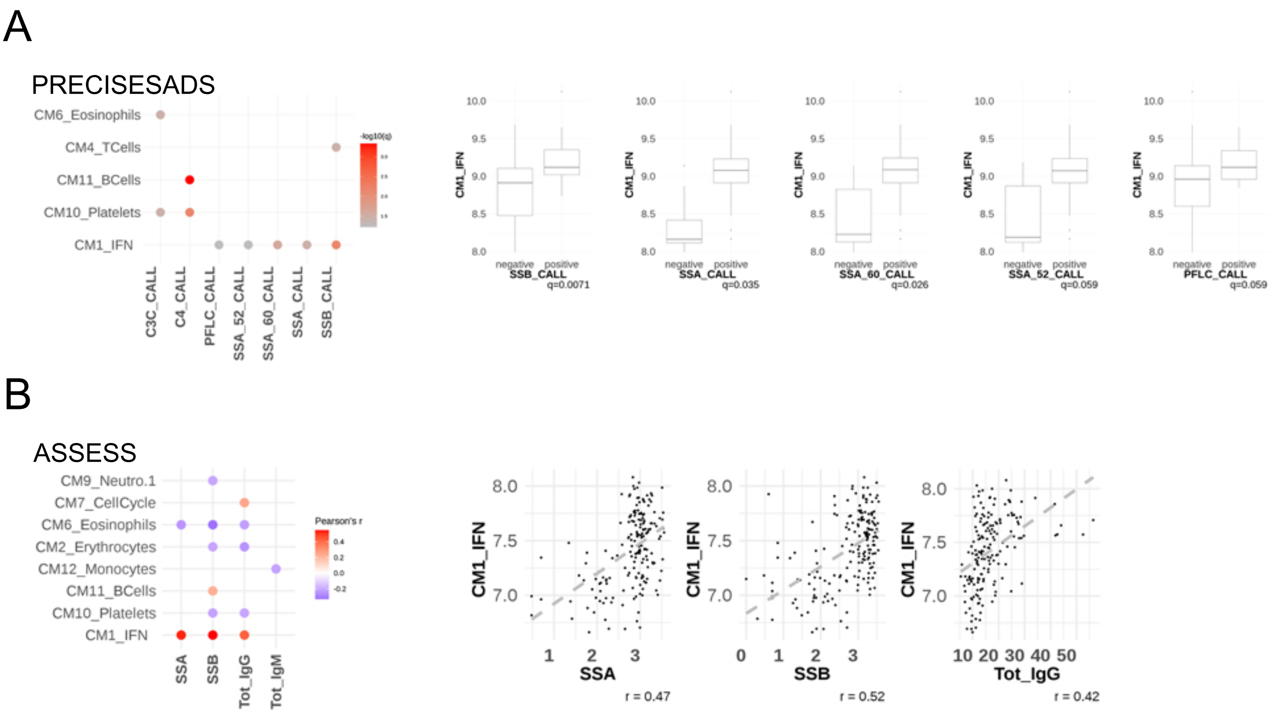


**Supplementary Fig5.** Pearson's correlation between average CMs expression and autoantibodies levels in A) PRECISESADS and B) ASSESS cohorts

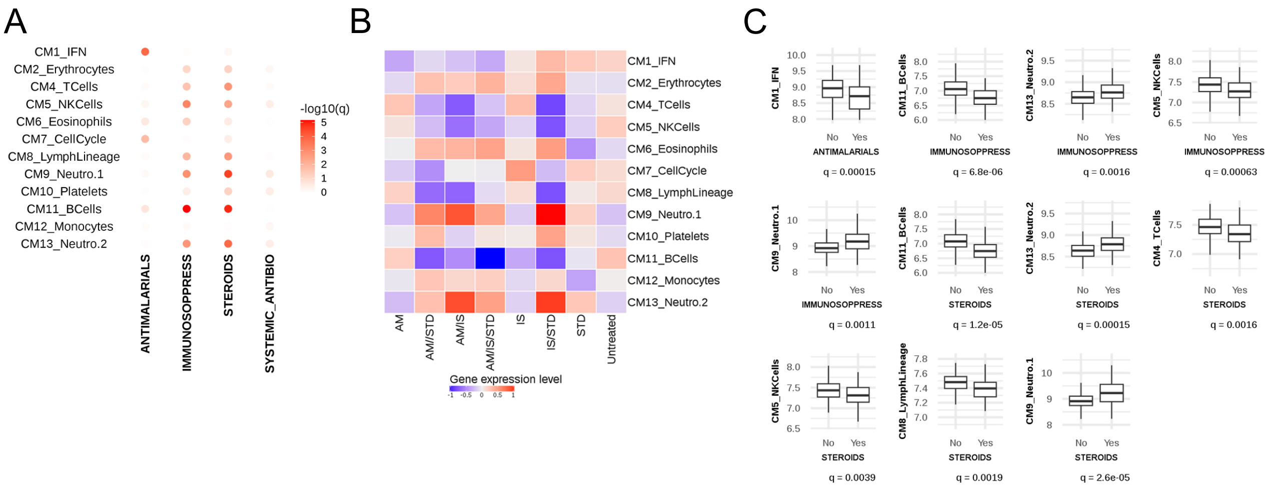


**Supplementary Fig6.** **A)**T-test between average CMs expression and treatment. q = corrected p.value **B)**CMs expression scores across patients stratified by treatments received. AM = Antimalarials, STD = Steroids, IS = Immunosupressors **C)**Significant diffrences observed in treated versus untreated patients.

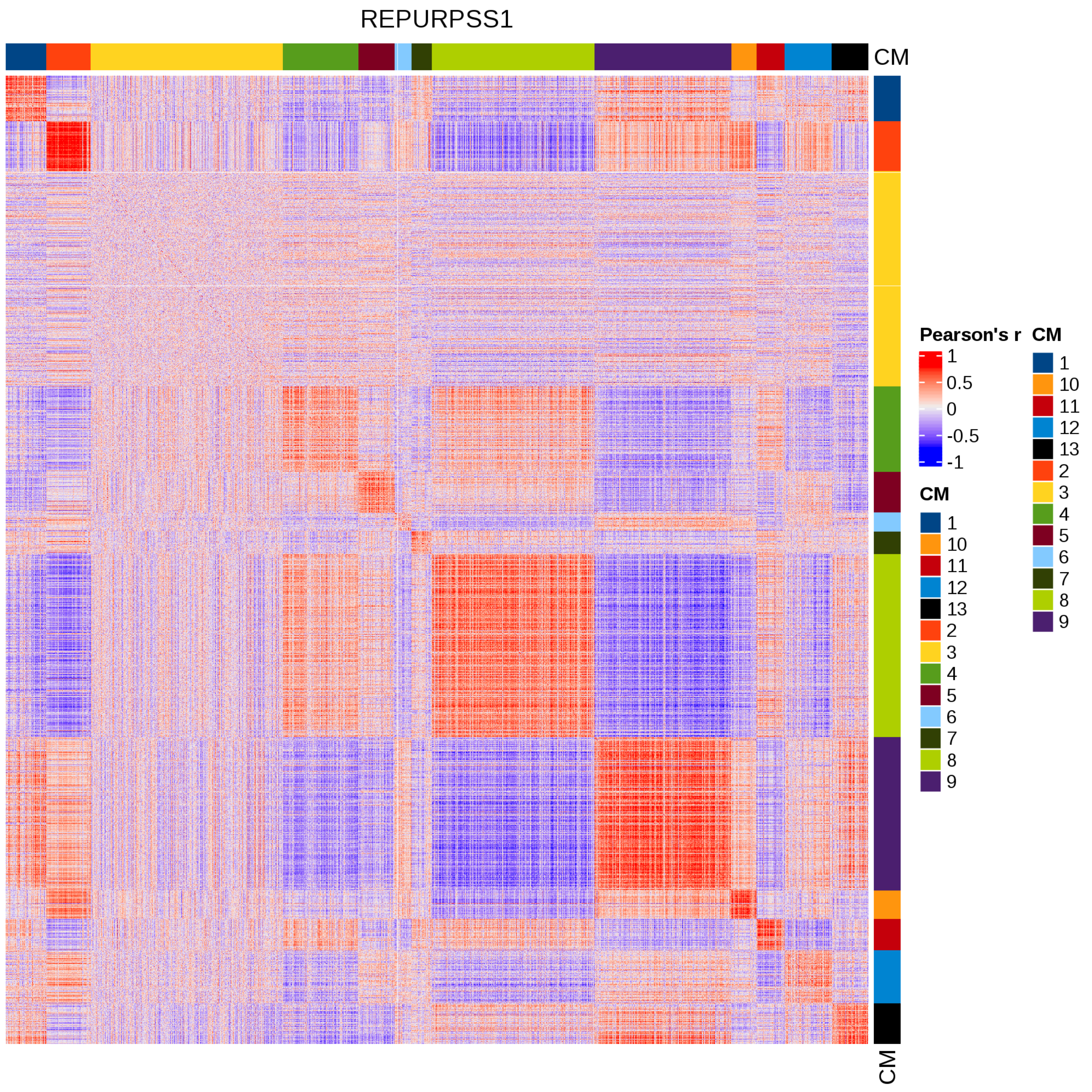


**Supplementary Fig7.** Correlation matrix in REPURPSS-1 cohort, sorted by CMs.

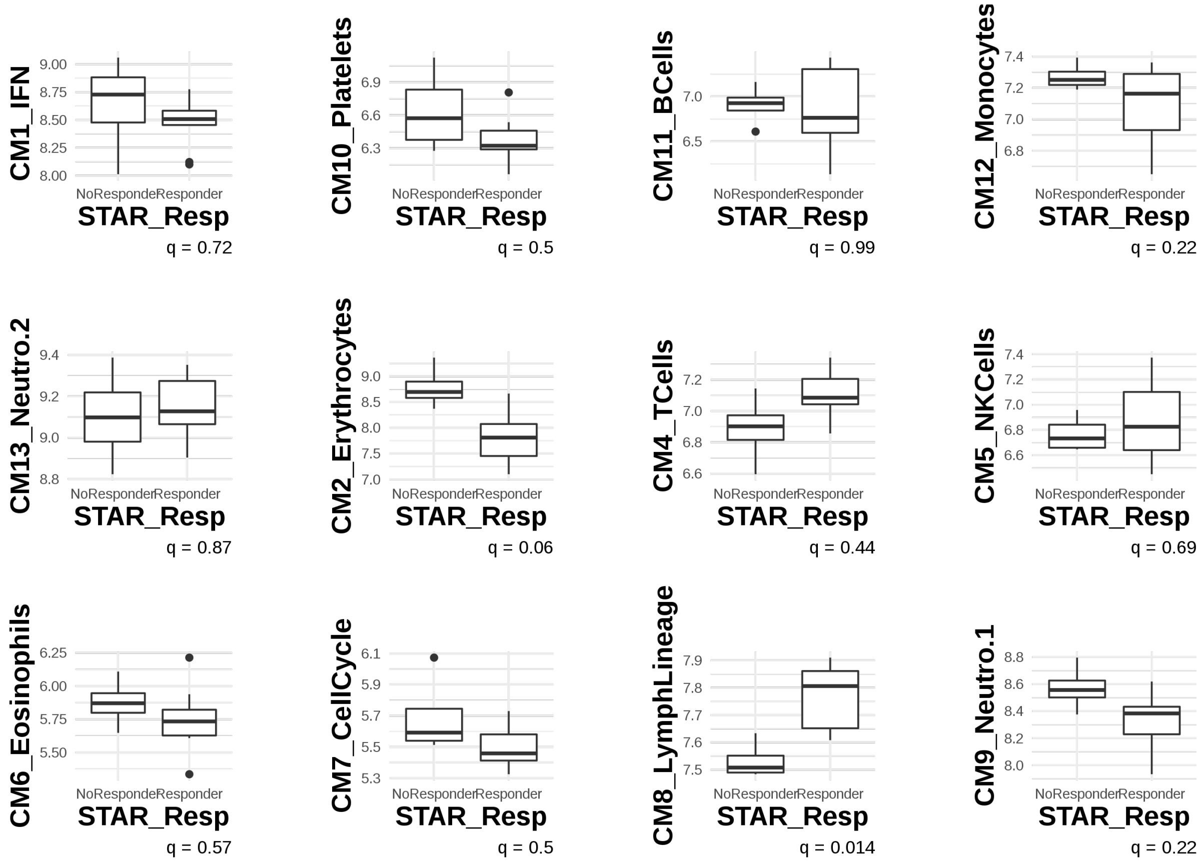


**Supplementary Fig8.** Boxplots of average expression of the CMs versus response status.
